# Improving Cycle Corrections in Discrete Time Markov Models: A Gaussian Quadrature Approach

**DOI:** 10.1101/2020.07.27.20162651

**Authors:** Tushar Srivastava, Mark Strong, Matthew D Stevenson, Peter J Dodd

## Abstract

**Introduction:** Discrete-time Markov models are widely used within health economic modelling. Analyses usually associate costs and health outcomes with health states and calculate totals for each decision option over some timeframe. Frequently, a correction method (e.g. half-cycle correction) is applied to unadjusted model outputs to yield an approximation to an assumed underlying continuous-time Markov model. In this study, we introduce a novel approximation method based on Gaussian Quadrature (GQ).

**Methods:** We exploited analytical results for time-homogeneous Markov chains to derive a new GQ-based approximation, which is applied to an unadjusted discrete-time model output. The GQ method approximates a continuous-time Markov model result by approximating a correction matrix, formulated as an integral, using a weighted sum of integrand values at specified points. GQ approximations can be made arbitrarily accurate by increasing ‘order’ of the approximation. We compared the first five orders of GQ approximation with four existing cycle correction methods (half-cycle correction, trapezoidal and Simpson’s 1/3 and 3/8 rules) across 100,000 randomly generated input parameter-sets.

**Results:** We show that first-order GQ method is identical to half-cycle correction method, which is itself equivalent to trapezoidal method. The second-order GQ is identical to Simpson’s 1/3 method. The third, fourth and fifth order GQ methods are novel in this context and provide increasingly accurate approximations to the output of the continuous-time model. In our simulation study, fifth-order GQ method outperformed other existing methods in over 99.8% of simulations. Of the existing methods, Simpson’s 1/3 rule performed the best.

**Conclusion:** Our novel GQ-based approximation outperforms other cycle correction methods for time-homogeneous models. The method is easy to implement, and R code and an Excel workbook are provided as supplementary materials.

## 1 Introduction

Discrete-time state-transition Markov models are common in healthcare decision analytical modelling. A discrete-time Markov model assumes that transitions between health states in a disease or treatment process occur at fixed intervals [1,2], but in reality, transitions between health states can usually occur at any point in time. A *continuous*-time Markov model is therefore a more natural representation for most disease and treatment processes. [1–3] Forcing transitions to occur only at discrete time steps leads to biased predictions of cumulative outcomes (e.g. cumulative costs, or cumulative utilities), and it is common for some kind of correction method to be applied. A correction method (e.g. the half-cycle correction [HCC]), applied to the cumulative outcomes of an unadjusted discrete-time state-transition Markov model, will result in model output that is closer to that which would have been observed in the continuous time analogue. [1–4]

There are various cycle correction method that exist, [5] the most common being the half-cycle correction method. [3, 6] This method assumes that, on average, transitions occur *halfway* through each time step, rather than at the beginning or end of each cycle, and simply subtracts and adds half of the outcome in each health state from the first and last cycle, respectively (note that we use the terms ‘cycle’ and ‘time step’ interchangeably.) [3, 6] Other methods from the numerical integration literature have also been used in decision analytical models for cycle correction. [5] These methods include the trapezoidal rule, Simpson’s 1/3 rule, and Simpson’s 3/8 rule, which all apply corrections at each cycle, not only at the first and last cycle. [5] Recently, Elbasha and Chhatwal (2016) reported that Simpson’s 1/3 method has the better accuracy compared to trapezoidal and Simpson 3/8 method. [5]

In this paper, we propose a new approach for cycle correction based on Gaussian quadrature (GQ). Gaussian quadrature is a method for approximating the integral of a function using a weighted sum of evalutions of the function at carefully chosen points. [7] We show that a correction factor, which adjusts a discrete-time model so that it matches its ‘true’ continuous time analogue, can be expressed as an integral, and this integral can be approximated using Gaussian quadrature.

The structure of the paper is as follows: in the next section we describe the relationship between a discrete-time and an assumed underlying continuous-time Markov model. We then review four existing correction methods: the half-cycle correction method, the trapezoidal rule, Simpson’s 1/3 rule, and Simpson’s 3/8 rule. Our novel correction method based on Gaussian Quadrature is then introduced, along with a simulation study to demonstrate its performance relative to existing methods. We conclude with a short discussion and conclusions. Supplementary material to implement the GQ approximation in Microsoft Excel and in R can be downloaded from the journal website.

## 2 Discrete- and continuous-time Markov models

### 2.1 The discrete-time Markov model

We start by introducing a simple illustrative cohort Markov model with three health states: ‘well’, ‘unwell’ and ‘dead’. At each time step, well patients can remain well, move to the unwell state, or die. Patients in the unwell state can recover to the well state, remain unwell, or die. Dead is an absorbing state of the model. The probabilities of transition between states are described by transition matrix,

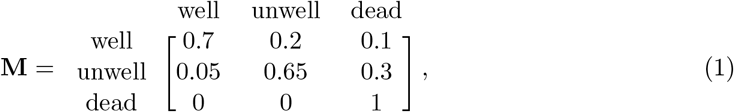

As the population cohort is assumed to be closed, each row of transition matrix sums to 1. We assume that the proportion of individuals in each health state at time zero is given by the row vector 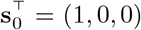, i.e. all individuals start in the ‘well’ state. The proportion of individuals in each health state at time step 1 is then given by the matrix product of 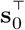 by **M**,

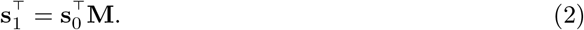

The proportion of individuals in each health state at time step *t* is found by repeated *matrix* multiplication of **M**,

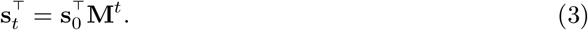

Note that **M**^*t*^ is not equal to the matrix obtained by raising the elements of **M** to the *t*-th power. In our example, health states are associated with the following per-time step costs (**c**) and utilities (**u**),

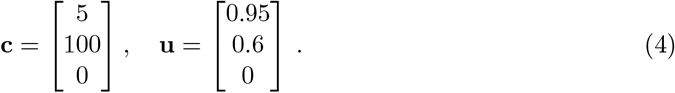

The total cost is determined by summing the costs associated with each time step over some predefined number of time steps (the ‘time horizon’), and similarly for total utility. In our example, the time horizon is 100 time steps. For brevity, we present methods and results for costs only. All arguments and expressions apply equally to utilities, simply by replacing **c** with **u**.

The cost accrued at time step *t*, denoted *C*(*t*), is given by

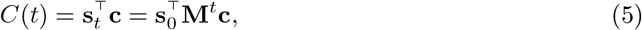

and the total cost over the period of time from *t* = 0 to *t* = *N* is given by

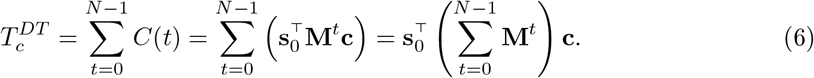

The superscript *DT* on the total costs *T*_*c*_ denotes that it is derived from the discrete-time model. In the discrete-time model as formulated in Equation (6), costs are assumed to occur at the *start* of each time period, hence *t* runs from 0 to *N* −1. If we assumed instead that outcomes occurred at the *end* of each time period, total costs would be given by 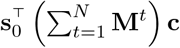, where *t* now runs from 1 to *N*.

See Figure 1(a) for a graphical illustration of how total costs are computed in a hypothetical discrete-time model where costs are assumed to occur at the start of each time period.

**Figure 1:**
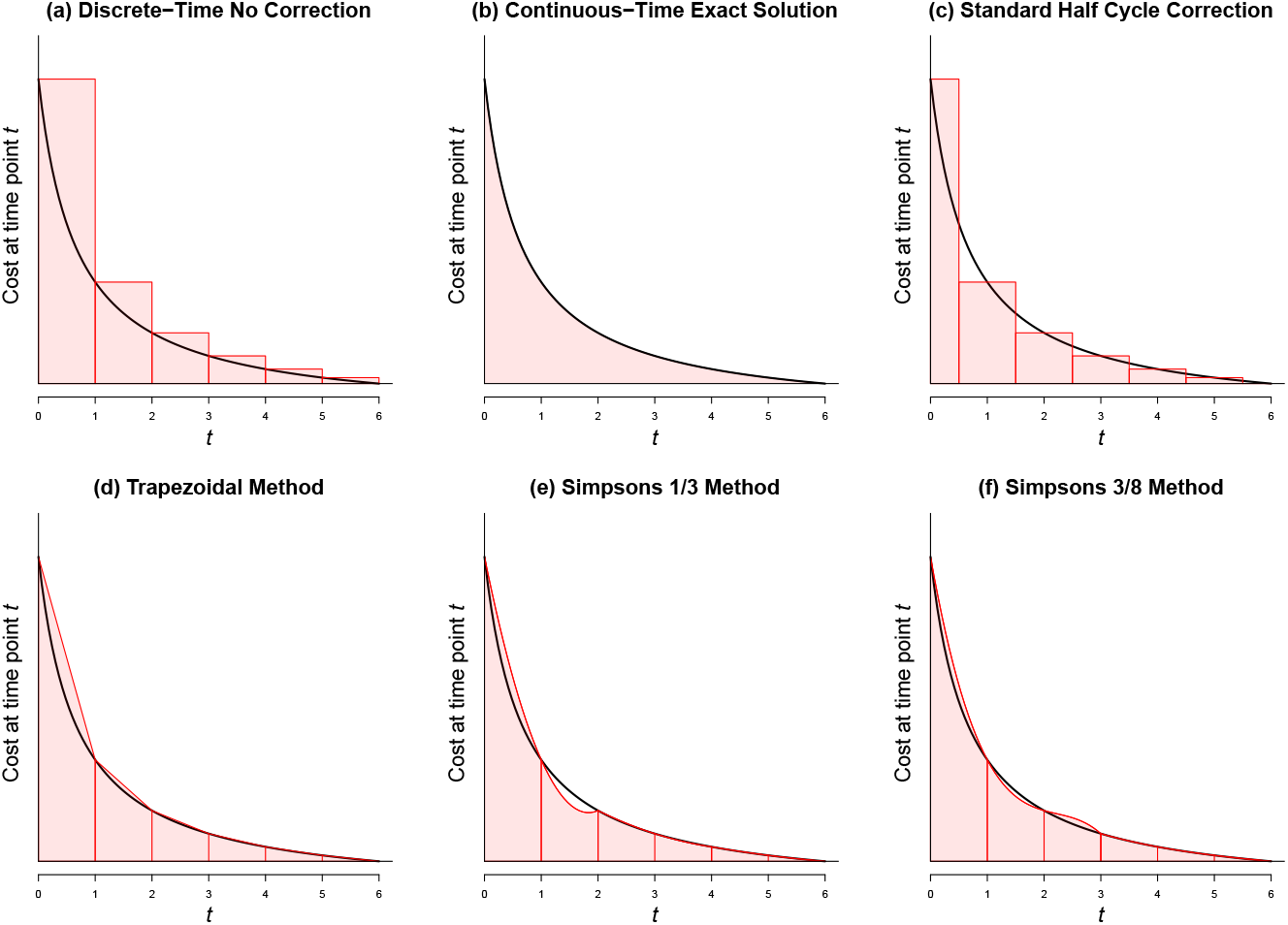
Graphical illustration of the (a) discrete- and (b) continuous-time models, and four correction methods (c-f) for the discrete-time model. The areas shaded in red represent total costs over a time horizon from t = 0 to t = 6.

Discrete-time Markov models are attractive due to their computational simplicity. However, in the health economic evaluation context in which disease processes are modelled, a discrete-time model will only approximate what is, in reality, a continuous process. We now introduce the continuous-time Markov model.

### 2.2 The continuous-time Markov model

Markov models can also be defined in continuous time via the Kolmogorov Forward Equation, a differential equation of the form

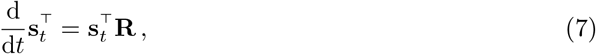

where 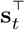 is a vector containing the proportion of individuals in each health states at time *t*, and **R** is a matrix of transition *rates* (sometimes also called the generator or intensity matrix). [8, 9] When **R** is constant (i.e. where rates are time-homogeneous), the evolution from the initial state to time *t* can be written in terms of a matrix exponential as

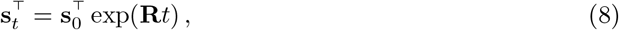

where 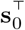 is the row vector containing the proportion of individuals in each health state at time zero.

Every time-homogeneous continuous-time Markov model gives rise to a discrete-time Markov model that, for unit time step, has transition probability matrix **M** = exp(**R**), where, again, exp() denotes the matrix exponential. For our model, the rate matrix that corresponds to transition probability matrix **M** is

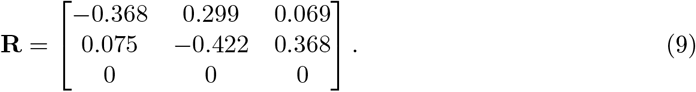

We can verify that **M** = exp(**R**) using the expm() matrix exponential function in the R package expm, and there are formulae for computing the matrix exponential by hand in low dimensions. [10] A discrete-event simulation for this continuous-time Markov chain could be implemented using 0.299 as the hazard of progressing from ‘well’ to ‘unwell’, 0.069 as the hazard of progressing from ‘well’ to ‘dead’, and so on.

In transition rate matrix **R**, rows sum to zero. This corresponds to the rows of **M** summing to 1, and indicates a closed cohort. The third row of zeroes corresponds to the absorbing state ‘dead’, which is not left once entered, and where there is no flow between states.

The cost at time *t*, denoted *C*(*t*), is given by

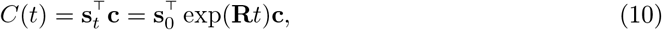

and the total cost over the time period from *t* = 0 to *t* = *N* is given by the integral

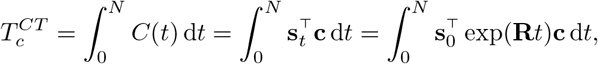

where the superscript *CT* on the total costs *T*_*c*_ denotes that it is derived from the continuous-time model. We can compute this integral analytically via the basic rules of integration, giving

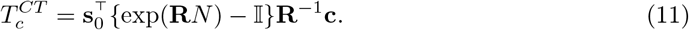

This is illustrated in Figure 1(b).

We note here that because the rate matrix, **R**, has rows that sum to zero, it will not have a full set of linearly independent columns, and will therefore not be invertible. However, equation (11) can still be evaluated if we replace the usual inverse with the *Moore-Penrose generalised inverse*. [11] This is implemented as the `ginv()` function in the `MASS` package in R.

### 3 Existing methods to correct a discrete-time model output

As we have noted, a discrete-time model is likely to be an approximation to a ‘true’ underlying continuous-time process. A discrete-time model assumes that transitions occur *at the time step*. No events occur *between* time steps because in a discrete time model, there is no time between steps; time is not continuous. In reality, in most settings, transitions between disease states can occur at any point in time. Therefore, an adjustment must be made to the output of a discrete-time model to address the bias that arises from the approximation.

### 3.1 Half-Cycle correction method

The ‘Half-Cycle’ correction (HCC) method is commonly used in health economic modelling. HCC assumes that the transition of the patient from one health state to another occurs midway between time steps, rather than at the time step. It is computed by subtracting and adding half of the outcome in the first and last cycle of the discrete-time model (equation 6), respectively, giving

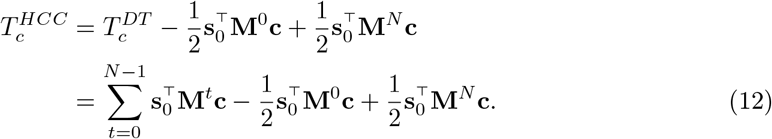

This is illustrated in Figure 1(c).

### 3.2 Trapezoidal method

This numerical integration technique, also commonly employed in health economic modelling, approximates an integral by dividing the range over which the function is integrated into trapeziums, and summing the areas of those trapeziums, [12] as illustrated in Figure 1(d).

Given two adjacent cycles, *t* and *t* + 1, with corresponding costs 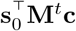 and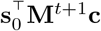, the area of the trapezium over this interval is given by

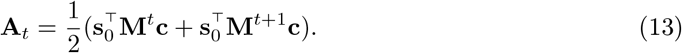

The total cost over the time horizon *N* is then given by:

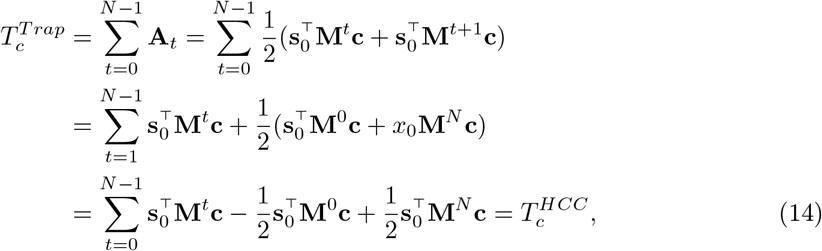

which, as shown in equation (14), is exactly equal to that given by half cycle correction method. The trapezoidal method is illustrated in Figure 1(d).

### 3.3 Simpson’s composite methods

Simpson’s composite methods uses a higher order polynomial function to connect adjacent points on the cost curve, rather than using a straight-line segment as used in the trapezoidal method. There are two commonly used types of Simpson’s rule: a quadratic approximation-based rule (Simpson’s 1/3), and a cubic approximation-based rule (Simpson’s 3/8) [13, 14]

#### 3.3.1 Simpson’s 1/3 rule

Given the costs computed for three adjacent cycles 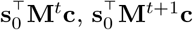 and 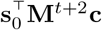, Simpson’s rule computes the area under a section of quadratic that passes through the three points. This is repeated then for the next three cycles 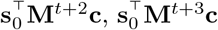 and 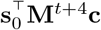, and so on.

The total cost can be shown to be

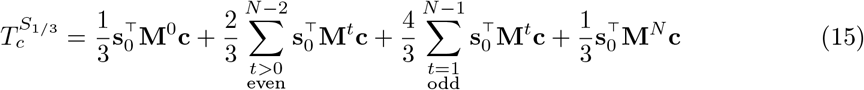

The Simpson’s 1/3 correction can be obtained by multiplying the outcomes by 1/3 in the first and last cycle and by 4/3 if the cycle number is odd and by 2/3 if the cycle number is even. Simpson’s 1/3 rule is illustrated in Figure 1(e). Note that in this method the time horizon *N* must be even.

#### 3.2.2 Simpson’s 3/8 rule

Similar to Simpson’s 1/3 rule, Simpson’s 3/8 rule uses sections of polynomial, but this time, cubic polynomial fitted to four adjacent points.

The total cost can be shown to be

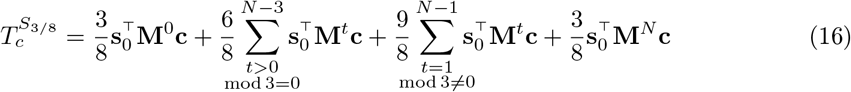

The Simpson’s 3/8 rule correction can be obtained by by multiplying the outcomes by 3/8 in the first and last cycle, by 6/8 if the cycle number is a multiple of 3 and by 9/8 otherwise. Simpson’s 3/8 rule is illustrated in Figure 1(f). Note that in this method the time horizon *N* must be an multiple of three.

## 4. New cycle correction method based on Gaussian quadrature

Our approach is to consider the difference between the total costs from the discrete- and continuous-time models, and attempt to find an approximation to this difference. Firstly, we derive an analytic expression for the total costs for a discrete-time model (6) by recognising that it is the sum of a finite geometric progression. For a *scalar, α*, the sum of a finite geometric progression is given by

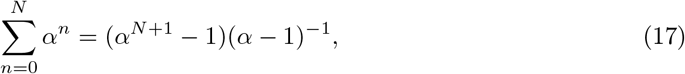

for any *α* ≠ 1. The matrix analogue for (17) is given by

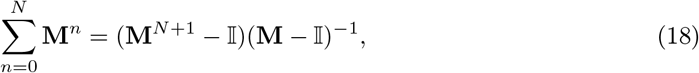

where 𝕀 is the identity matrix (in this case of dimension 3), and hence we can write the discrete-time model as

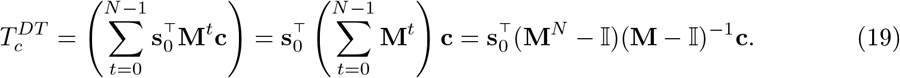

As an aside, this formula is only valid when (**M** − 𝕀) is invertible, which will *not* be the case if **M** is a transition matrix. This is because **M** − 𝕀 will have rows that sum to zero, and therefore will not have a full set of linearly independent columns. However, we can get around this problem, as long as the costs and utilities are zero for absorbing states (‘dead’ in our example), by replacing the usual matrix inverse with the Moore-Penrose generalised inverse (see Appendix for proof). [11, 15]

We now consider how we could adjust the discrete-time model so that the cumulative sum of outcomes equal those of the continuous-time model. Recall that the rate and transition matrices are linked via **M** = exp(**R**) and hence **M**^*N*^ = exp(**R***N*). We can therefore get to the continuous-time model cumulative sum of costs

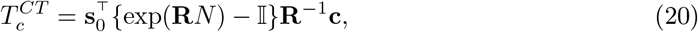

from the discrete-time model cumulative sum of costs

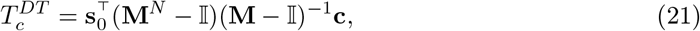

by inserting a correction factor of (**M** − 𝕀) log(**M**)^−1^ before **c** in the expression (21), as follows

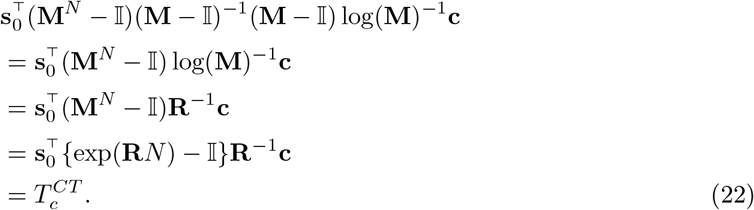

Recall that the Markov trace is the matrix that records the health state occupancy over time. It has the same number of columns as there are health states, and the same number of rows as there are time cycles. In our example, the matrix is of size 100 *×* 3. Summing down the columns of the Markov trace gives us 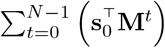, which we showed above can be written 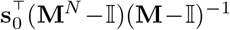. So, in practice, we simply need to post-multiply the sum of the columns of the Markov trace from our discrete time model by the correction factor (**M**− 𝕀) log(**M**)^−1^ before we then multiply by the cost vector **c**, i.e. replacing 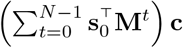 with 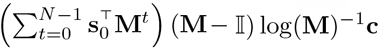.

We now turn to the question of how we find (**M** − 𝕀) log(**M**)^−1^. If we are using a coding language (R, Matlab, Python, etc) then we can evaluate (**M** − 𝕀) log(**M**)^−1^ directly using inbuilt matrix algebra functions. In R the correction factor would be (M-I) %*% `ginv(logm(M))`, where `ginv()` is the Moore-Penrose inverse function from the `MASS` package, `logm()` is the matrix log function from the `expm` package, and `I` is defined as the identity matrix of dimension *n* matching the dimensionality of **M**, via I <-`diag(nrow(M))`. If the sum of the columns of the Markov trace is denoted × in R, and our cost vector by c, then we can recover the continuous-time model total costs via × %*% (M-I) %*% ginv(logm(M)) %*% c.

This is trivial in R, but unfortunately, the matrix logarithm is very difficult to compute in a typical spreadsheet application such as Microsoft Excel, and this problem motivates the final step: the Gaussian quadrature approximation.

Firstly, we define **Z** to be the *inverse* of the correction factor (we will invert it back again later),

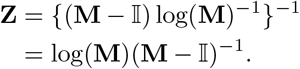

Next, we note that **Z** has an integral representation as

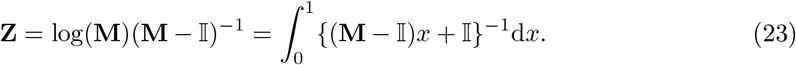

The proof of this is somewhat involved, and we refer interested readers to Wouk (1965) [16].

Finally, we develop an approximation to the integral representation of **Z** using Gaussian quadrature, in such a way that we can compute the approximation in a spreadsheet. Gaussian quadrature is a method for approximating the integral of a function by a sum of carefully chosen polynomials. In practice, this involves evaluating the function to be integrated at a set of pre-specified points, and summing the values with pre-specified weights. The points and weights can be found in standard tables. The number of terms in the summation is called the *order* of the GQ approximation, with higher orders giving better approximations.

Several Gaussian quadrature ‘rules’ (i.e. methods for determining the points and weights) exist, including *Gauss-Legendre* quadrature, which approximates the integral representation of **Z** in Equation 23 as

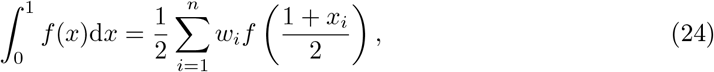

where *f* (*x*) = {(**M** − 𝕀)*x* + 𝕀}−1, the function to be integrated.

The locations {*x*_*i*_} and weights *{w*_*i*_*}* for first 5 orders (values of *n*) are given in Table 1.

**Table 1:**
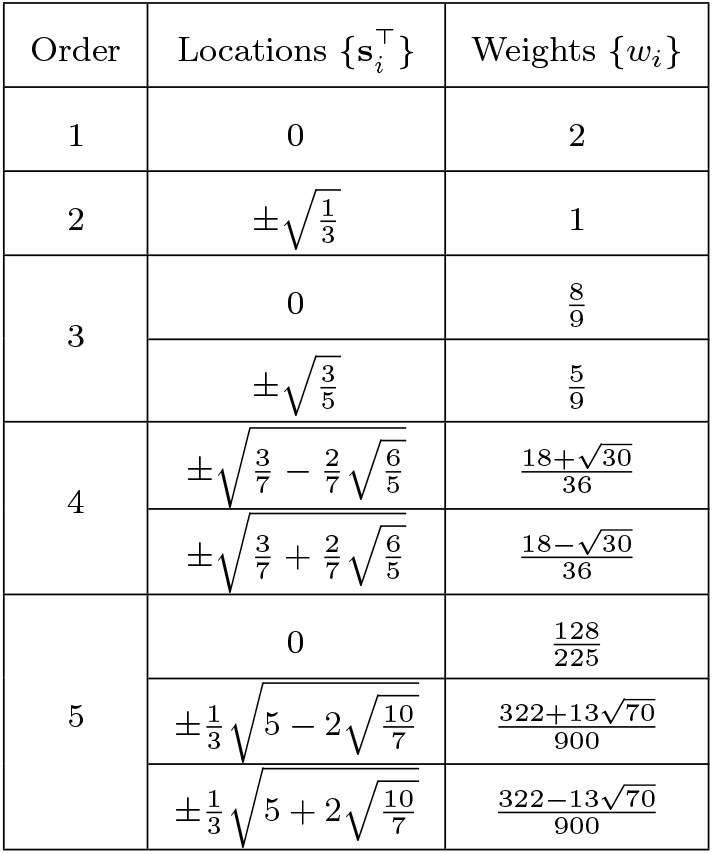
Locations and weights for the first 5 orders of Gaussian-Legendre quadrature.

Plugging these values into Equation 24, gives the following approximations for **Z** at orders *n* = 1 and 2:

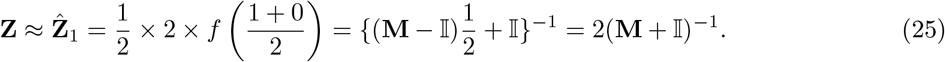

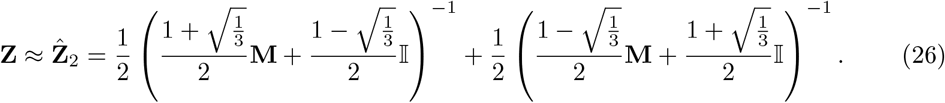

Orders 3, 4 and 5 can be calculated similarly by evaluating *f* (*x*) at the locations in Table 1, and summing with the relevant weights. Fully-worked formulae are also in the supplementary material.

To summarise, we run the unadjusted discrete-time Markov model to generate the Markov trace, sum down the columns of the Markov trace, post-multiply this by the inverse of 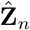 (where *n* is the chosen order for the GQ correction), and finally post-multiply the result by the cost vector **c**.

The first order GQ approximation is equivalent to the trapezoidal method (which, as we have shown, is equivalent to the half-cycle correction) This can be shown as follows:

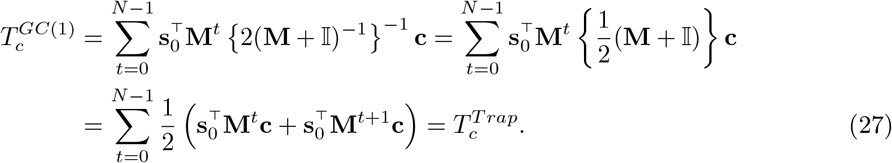

The second order GQ method can be shown to be identical to Simpson’s 1/3 method, but the third order GQ method is novel and should result in a better approximation than either the half cycle or Simpson’s 1/3 methods. Higher order GQ methods will results in better approximations still.

## 4.1 Discounting

Normally, analyses in health economics employ discounting. For discrete-time models, we multiply costs accrued at time step *t* by a factor 1/(1 + *ρ*)^*t*^, where *ρ* is the per-time-step discount rate. This is equivalent to replacing the transition probability matrix **M** with **M**^*∗*^ = **M**/(1 + *ρ*),

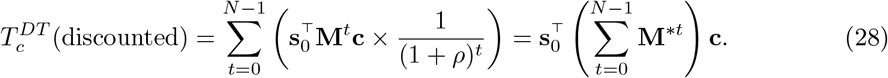

For a continuous-time model, we would add the natural logarithm of the discount factor *ρ*^*∗*^ = log*{*1/(1 + *ρ*)*}* to the diagonal elements of **R**, giving **R**^*∗*^ = **R** + *ρ*^*∗*^𝕀, and therefore

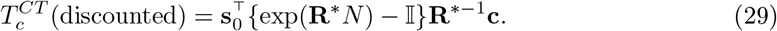

All the approximation methods we have described in this section hold equally when discounting is employed and we will not consider this further, other than to note that (**M**^*∗*^ − 𝕀) and **R**^*∗*^ are both invertible, and we therefore do not need to use the Moore-Penrose generalised inverse when outcomes are discounted.

## 5 Simulation Study

We first undertook a deterministic analysis in which we assumed that the transition matrix **M**, cost vector **c**, utility vector **u** and starting health state proportion vector 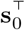 took, with certainty, their values as presented above.

We then performed a probabilistic sensitivity analysis (PSA) to explore the impact of uncertainty on input parameters of the model. [17] In this analysis, transition probabilities, costs and utilities were each assigned a probability distribution. For costs, we assumed lognormal distributions, for utilities, we assumed beta distributions, and for the rates in matrix **R** we assumed exponential distributions (Table 2). The transition probability matrix **M** was derived by taking the matrix exponential of the rate matrix *R*. All parameters were considered independent. See supplemental Figure S4 for histograms of these distributions.

**Table 2:** Model parameters with mean values, standard deviation and probabilistic distribution used for PSA. Rate rij is the entry in the i^th^ row and j^th^ column of the rate matrix **R**. Abbreviations - PSA: Probabilistic Sensitivity Analysis

| Model Inputs | Mean Value | Standard Deviation | Distribution Used in PSA |
| --- | --- | --- | --- |
| Well State Cost | £5 | £1 | Log Normal (1.6,0.198) |
| Unwell State Cost | £100 | £20 | Log Normal (4.61,0.198) |
| Dead State Cost | £0 | £0 | Assumed zero with certainty |
| Well State Utility | 0.95 | 0.19 | Beta (0.3,0.016) |
| Unwell State Utility | 0.6 | 0.12 | Beta (9.4,6.27) |
| Dead State Utility | 0 | 0 | Assumed zero with certainty |
| $r_{12}$ (Well to unwell rate) | 0.299 | 0.299 | Exp(1/0.299) |
| $r_{13}$ (Well to dead rate) | 0.069 | 0.069 | Exp(1/0.069) |
| $r_{21}$ (Unwell to well rate) | 0.075 | 0.075 | Exp(1/0.075) |
| $r_{23}$ (Unwell to dead rate) | 0.368 | 0.368 | Exp(1/0.368) |
| $r_{31}$ (Dead to well rate) | 0 | 0 | Assumed zero with certainty |
| $r_{31}$ (Dead to unwell rate) | 0 | 0 | Assumed zero with certainty |
| $r_{11}$ (Well to well rate) | | | $-(r_{12} + r_{13})$ |
| $r_{22}$ (Unwell to unwell rate) | | | $-(r_{21} + r_{23})$ |
| $r_{33}$ (Dead to dead rate) | | | $-(r_{31} + r_{32})$ |

To ensure good coverage of parameter space, a Latin hypercube design was used to draw 100,000 parameter sets. For each parameter set, we computed the total costs and total QALYs for the discrete-time model, continuous-time model, and the six approximation methods: (i) half-cycle = trapezoidal = 1st order GQ; (ii) Simpson’s 1/3 rule = 2nd order GQ; (iii) Simpson’s 3/8 rule; (iv) 3rd order GQ; (v) 4th order GQ; and (vi) 5th order GQ. In each case, we computed the absolute relative error (i.e. the absolute value of the difference between the approximation and the continuous model output, divided by the continuous model output) for costs and for QALYs.

To understand the impact of cycle length on approximations, we also performed scenario analysis with varying cycle lengths from 1 week to 52 weeks.

All the analyses were performed in both Microsoft Excel and in R, and code can be found in the supplementary material.

## 6 Results

Results are presented in Table 3. In the deterministic analysis, the 3rd order GQ method outperformed existing methods, with higher order GQ approximations further improving the approximation. Simpson’s 1/3 method (which is identical to the 2nd order GQ method) was the best performing of the existing methods. For total costs in our example, the half-cycle correction method (equivalent to the trapezoidal and 1st order GQ methods) gave a worse approximation than no correction. In addition, the difference in net-monetray benefit (NMB) between our proposed method and the half-cycle method is over £400 which implies that the method employed for continuity correction could alter decision-making.

**Table 3:** Comparative analysis result per person based on different methods. Absolute error is presented in increasing order of error. 5th order GQ method has the absolute minimum error for both costs and QALYs and closest to continuous model. 1st order GQ is same as HCC and trapezoidal and 2nd order GQ is same as Simpson 1/3. NMB is with respect to continuous time model with willingness-to-pay of £30,000. Abbreviations: GQ: Gaussian Quadrature; HCC: Half-Cycle Correction; NMB: Net Monetary Benefit; QALY: Quality Adjusted Life-Years

| Correction Method | Costs | Absolute Error | QALYs | Absolute Error | NMB |
| --- | --- | --- | --- | --- | --- |
| Continuous time model | £228.7622 | - | 4.27397 | - | - |
| 5th order GQ method | £228.7622 | 1.86e-07 | 4.27397 | 6.85e-07 | -£0.0877 |
| 4th order GQ method | £228.7621 | 3.33e-07 | 4.27397 | 7.03e-07 | -£0.0900 |
| 3rd order GQ method | £228.7604 | 7.73e-06 | 4.27396 | 1.55e-06 | -£0.1972 |
| 2nd order GQ method | £228.6821 | 3.51e-04 | 4.27386 | 2.54e-05 | -£3.1738 |
| Simpson's 3/8 method | £228.5886 | 7.59e-04 | 4.27374 | 5.29e-05 | -£6.6033 |
| 1st order GQ method | £226.4474 | 1.01e-02 | 4.28816 | 3.32e-03 | £427.9517 |
| No correction performed | £228.9474 | 8.09e-04 | 4.76316 | 1.14e-01 | £14,675.4517 |

Results with discounting are presented in supplementary Table S1

The PSA results are presented in Figure 2. The box and whisker plot presents the absolute relative errors on the log scale for each correction method, compared against the continuous-time model output. The 5th order GQ method has the smallest median absolute relative errors, followed in increasing order of error by the 4th order GQ, 3rd order GQ and then Simpson’s 1/3 method.

**Figure 2:**
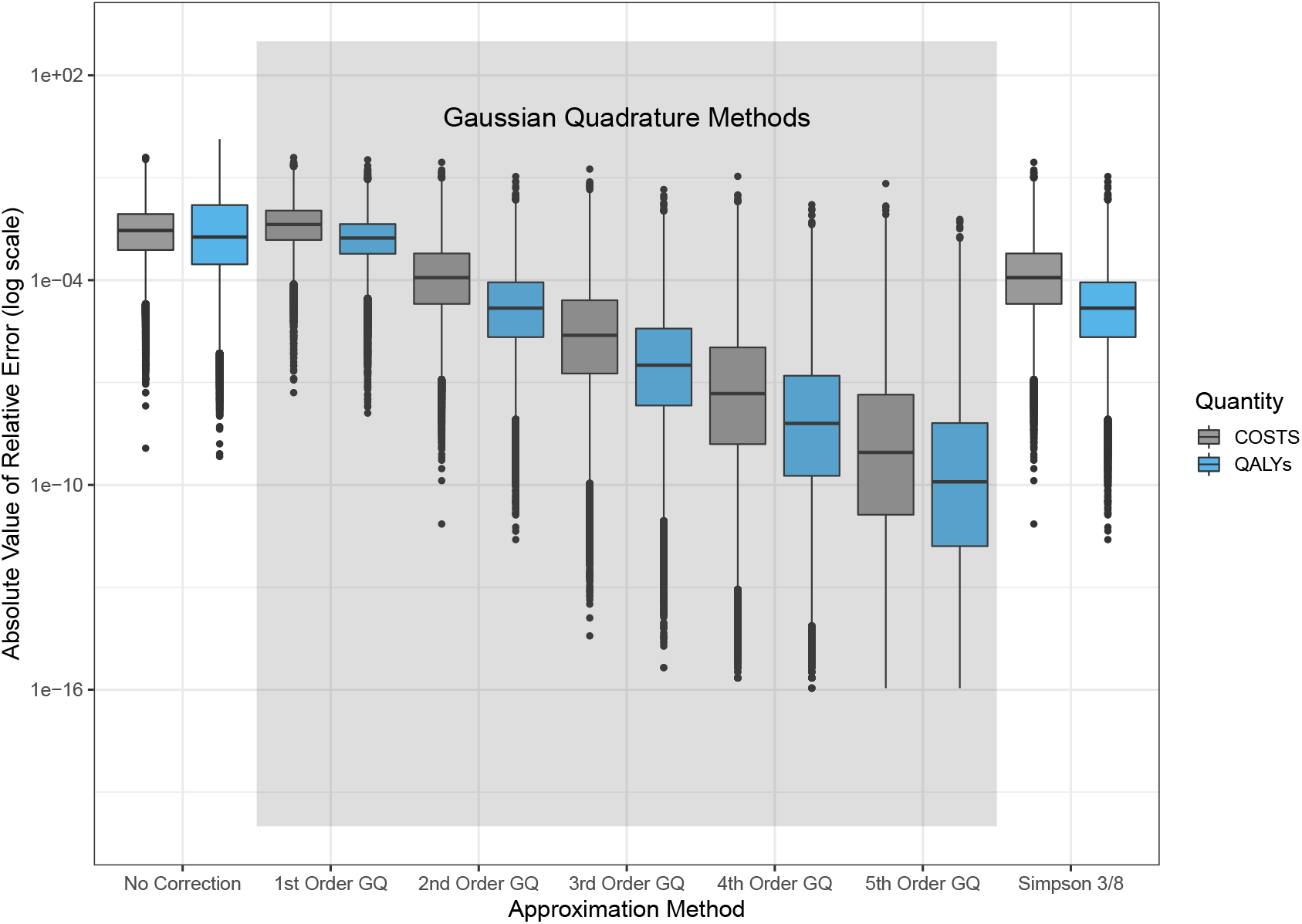
Probabilistic Sensitivity Analysis Result: Absolute value of the relative errors for cost and QALY in 100,000 PSA runs between the continuous time model and discrete time model with different cycle correction methods.1st order GQ is same as HCC and trapezoidal; 2nd order GQ is same as Simpson 1/3. Costs and QALYs outcomes corrected with 5th order GQ method has the minimum error. Abbreviations: GQ: Gaussian Quadrature; HCC: Half-Cycle Correction; QALY: Quality Adjusted Life-Years

A rankogram in Figure 3 shows that in 100,000 PSA runs, the 5th order GQ approximation method performed best in the 99.8% of cases (i.e. it was not ranked first in 0.2% of runs). Whereas, Simpson’s 1/3 rule (the best of the existing methods) ranked first in only 0.001% runs.

**Figure 3:**
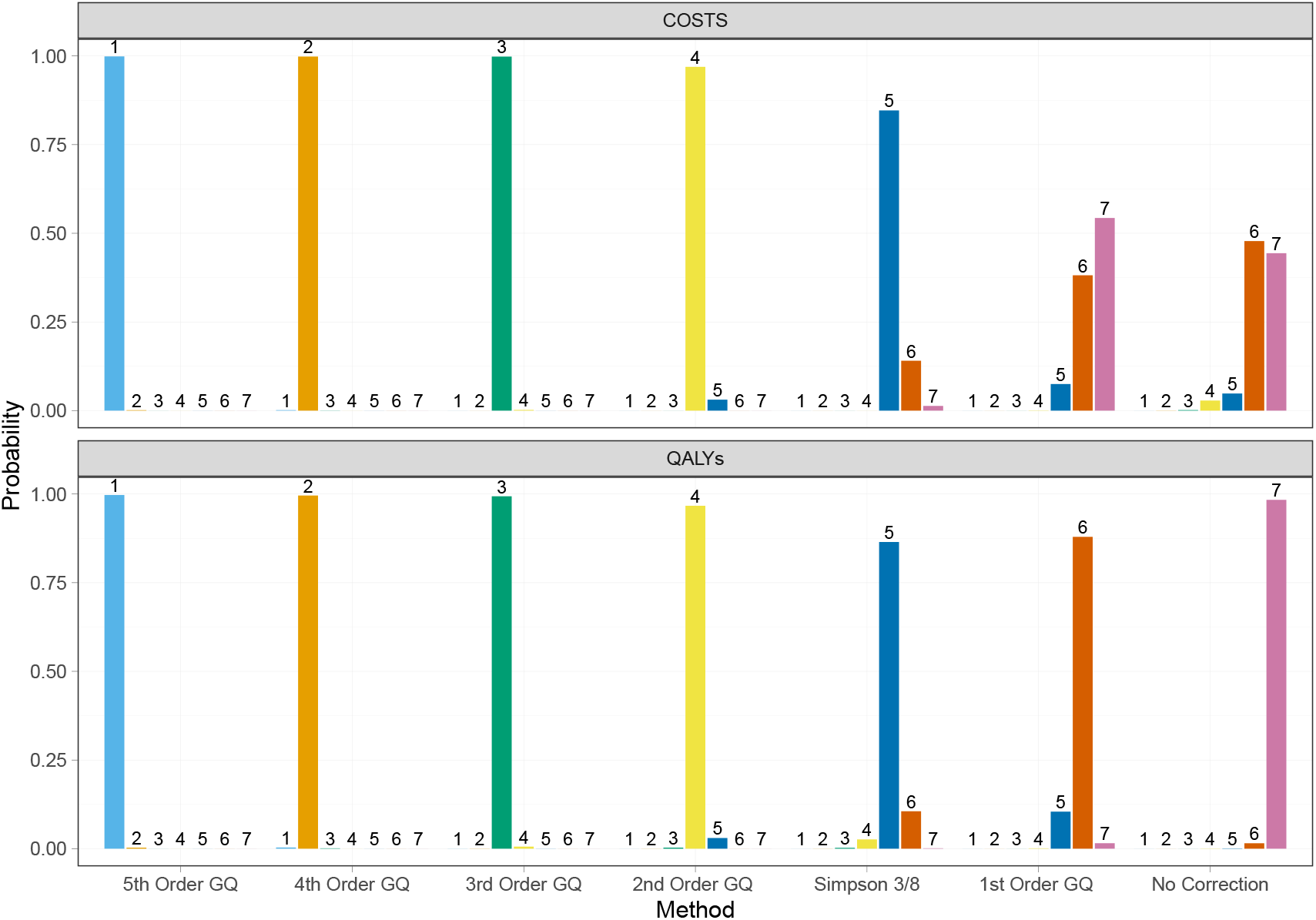
Rankogram bar plot showing ranks for accuracy (absolute value of relative error) of each approximation method in 100,000 PSA runs; where rank 1 represents minimum error and rank 7 maximum error. 1st order GQ is same as HCC and trapezoidal; 2nd order GQ is same as Simpson 1/3. Abbreviations: GQ: Gaussian Quadrature; HCC: Half-Cycle Correction; PSA: Probabilistic Sensitivity Analysis; QALY: Quality Adjusted Life-Years

When we examined the parameters of the runs in which the 5th order GQ approximation was not best, we discovered two patterns. Firstly, the 5th order GQ approximation suffered when the transition probabilities for well to well, and from unwell to unwell were both close to 1 (i.e. then transitions between states were rare, and **M** was close to the identity matrix). In these cases, all methods performed well, with the best being the 4th rather than the 5th order GQ method. However, the differences between the 4th and 5th order approximations were tiny, with absolute relative errors of the order 10^−14^, and therefore unlikely to be of any consequence. These runs form the vast majority of 5th-order GQ ‘failures’, and appear as a peak in the density plots shown in supplementary Figures S2 and S3. Secondly, the 5th order GQ approximation did less well when the probabilities for ‘forward’ transitions (i.e. from well to unwell, from well to dead, and from unwell to dead) were high, resulting in 100% mortality within a few cycles. Indeed, in this case, *all* of the correction methods performed poorly, with the uncorrected output being closest to the continuous-time model result. There were very few failures of this type (*<* 0.01% of all runs, and these failures are therefore not visible in supplementary Figures S2 and S3.)

The impact of cycle length on the quality of the approximations is presented in Figure 4. With a shorter cycle length, the absolute error between continuous- and discrete-time model using different approximation methods decreases as expected. The 5th order GQ method performs best overall.

**Figure 4:**
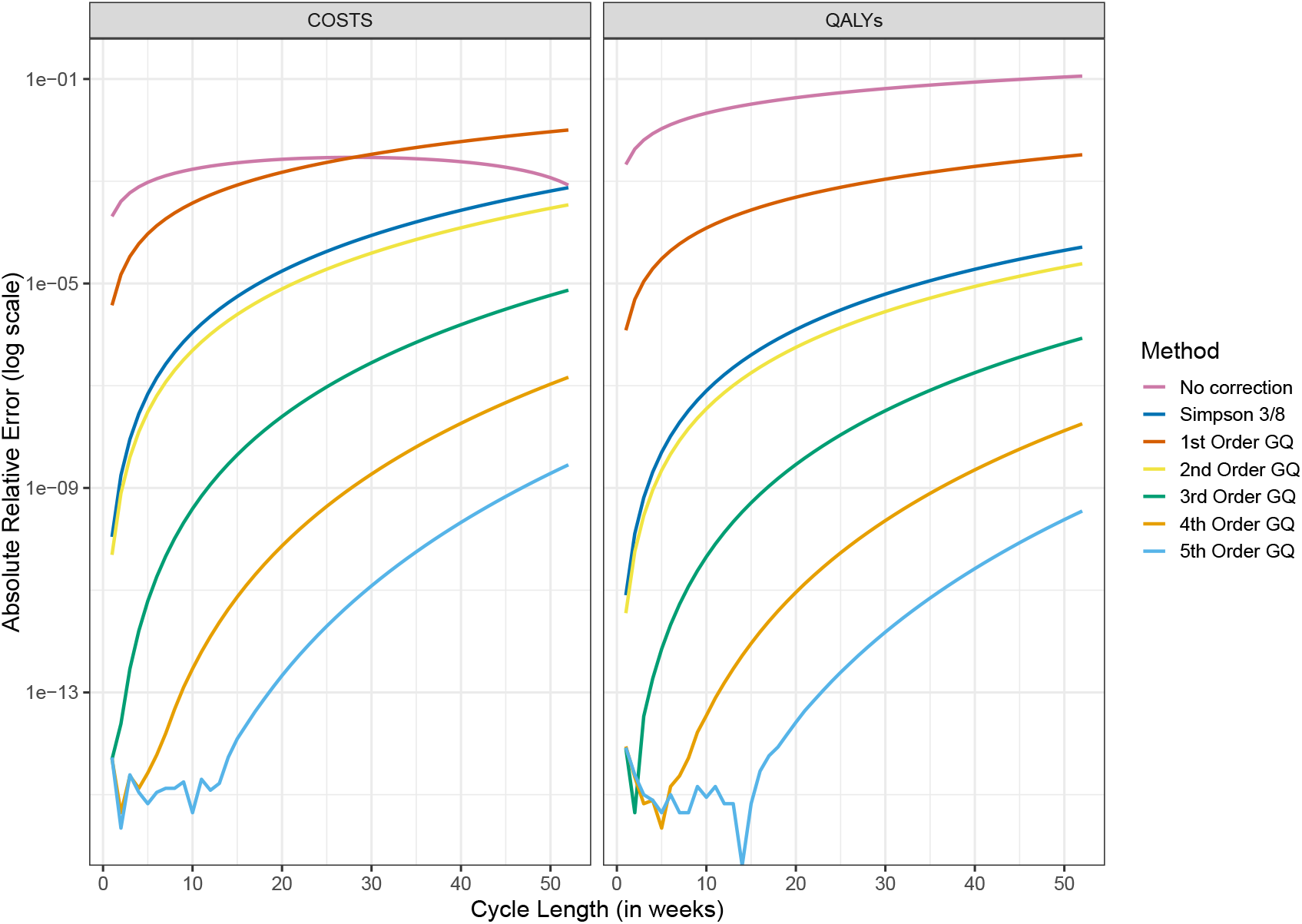
Impact of cycle length on absolute error between continuous- and discrete-time model using different approximation methods.1st order GQ is same as HCC and trapezoidal and 2nd order GQ is same as Simpson 1/3. Most of the time 5th order GQ method has lowest error compared to the other methods. Abbreviations: GQ: Gaussian Quadrature; HCC: Half-Cycle Correction; QALY: Quality Adjusted Life-Years

The impact of time horizon on the quality of approximation is presented in supplementary Figure S1 and similar results were observed. In each case, the 5th order GQ method had the lowest error.

## 7 Discussion

### 7.1 Main results

In our study, we found that the 3rd, 4th and 5th order GQ methods all outperformed existing cycle correction approximation methods, with higher orders giving better approximations. We chose the 5th as the highest order GQ to implement, but the adjusted discrete-time model output can be made arbitrarily close to the continuous-time model output by increasing the order of the GQ approximation. When we explored higher orders in our simulation study, we found very little benefit to orders greater than five.

For a discrete-time step Markov model, it is important to have a correction method which can be easily applied. Our GQ method can be applied as a post-hoc correction to an uncorrected Markov trace, and can be easily implemented in spreadsheet software such as Microsoft Excel. In previous comparisons of existing methods, Simpson’s 1/3 rule gave the best approximation to the continuous-time model, [5, 18] and our results support this finding.

In our simple numerical example, the standard half-cycle correction yielded a *worse* approximation than the uncorrected output from the discrete-time model. This is a concern, and suggests that half-cycle correction method should be avoided.

### 7.2 How this fits with the existing literature

There are a number of papers in the literature on correction methods for discrete time models, see, for example [5, 6, 18–20]. The main published methods are the half-cycle correction, the trapezoidal and Simpson’s methods, and the life-table method. We did not include the life-table method in our analysis because it has been shown to be equivalent to the trapezoidal method (and therefore also to the half-cycle method). [5, 18]

Elbasha and Chhatwal (2016) highlighted the need to test a range of other methods, including Gaussian quadrature, Simpson’s 6-points rule, Boole’s rule and Romberg’s quadrature rule. However, in all of the literature so far, the target for approximation has been the cumulative total outcome (costs and QALYs in our example). What is unique about our approach is that we use Gaussian quadrature to approximate a *correction factor* (that post-multiplies the Markov trace) rather than approximating the cumulative total outcome itself. Our simulation study suggests that even a low-order (i.e. 3rd-order) GQ approximation results in an excellent correction.

### 7.3 Strengths and limitations

The GQ method is easy to implement in both spreadsheet software and in R, and can be applied as a *post hoc* correction to uncorrected Markov traces. We provide an Excel spreadsheet and R implementations as supplementary materials. Extension to a higher orders than the 5th is straightforward, though the benefit is likely to be inconsequential.

Our study has limitations. We have not computed bounds for the relative errors, or established rigorous criteria for when the 5th order GQ is guaranteed to outperform other methods. Furthermore, although we undertook a PSA to investigate the robustness of our results to different parameters, we used only a single model. We did not investigate the effect of different state-space dimension on our approximation’s performance. The model type considered for the simulation study was one with the constant transition matrix rather than time-varying transition matrix. While it is not obvious how to generalize our approximation scheme directly to the time-varying case, use of tunnel states can approximate time dependence by increasing the dimension of the model state-space.

### 7.4 Problems with discrete-time models

Every rate matrix **R** gives rise to a unique transition probability matrix **M** for time step *t* via **M** = exp(**R***t*). This means that every continuous-time model can be expressed as a discrete-time model. However, the converse is not necessarily true. Given **M**, there may not be a unique rate matrix **R**, or **R** may not exist at all (see e.g. Higham [21]). Indeed, determining whether a transition matrix **M** has a unique rate matrix **R**, is an unsolved problem in probability theory, where it is known as the ‘embedding’ (or ‘embeddability’) problem. [22]

This presents a specific problem that arises in practice, namely determining the transition matrix for shorter cycle length than originally specified (eg. the monthly transition matrix for a model defined by a yearly transition matrix). The typical approach to this is to compute the *p*-th root of the matrix, where *p* is the multiplicative factor between the time periods (e.g. 12 for converting the yearly cycle model to the monthly cycle model). Even when it is defined (which is not guaranteed), the *p*-th root of **M** may not be a valid transition matrix. In the context of financial applications, methods have been developed that choose approximate *p*-th roots for models specified via transition matrices, [23, 24] however models specified in this way remain unsatisfactory.

All these issues are absent for models that are defined in terms of a rate matrix **R**; transition matrices over any time period are guaranteed to be valid, and are unique. This is an important motivation for modelling healthcare decision problems in terms of their continuous-time dynamics, rather than artificially as discrete-time systems, but the historical reliance on spreadsheet applications has rather hampered the adoption of continuous time models.

We would argue that defining models in continuous time is a more natural and logically consistent representation of reality, and obviates the need for correction methods or stochastic simulation. Often data on the efficacy of interventions are reported as hazard ratios, which can be applied more naturally to continuous-time models defined in terms of rates. Methods for estimating rates from discrete-time data are available, [9, 25] including as an R package. [26]

Finally, where cycle corrections are employed, we would caution against their unthinking use. Some costs may be periodic and aligned with cycle length, eg. injections with a set frequency. These costs are incurred by all relevant compliant patients at the start of a cycle; if cycle correction are performed, intervention costs will be underestimated.

## 7.5 Conclusion

A novel Gaussian quadrature-based method for correcting the output of a discrete-time Markov model provides a better approximation to an assumed underlying continuous-time Markov model than do current methods.

## Data Availability

Excel and R model are provided as a supplementary material

## Authors’ Note

This work was done in ScHARR, University of Sheffield, UK. An abstract from this work was presented as a poster presentation at the International Society of Pharmacoeconomics and Outcomes Research (ISPOR) - EU, Denmark in November 2019

## A Appendix: Justification for the use of the Moore-Penrose generalised inverse

We wish to establish a sufficient condition for

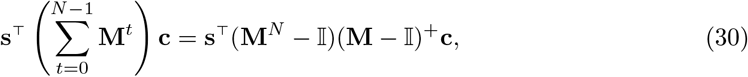

to be true.

As in the paper, we will write **A** = (**M** − 𝕀), and **A**^+^ for the Moore-Penrose inverse of **A**. We recognise that Equation 30 is valid if:

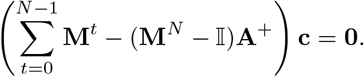

We can show this is true if we can show that **c** = **AA**^+^**c**. This is because, if **c** = **AA**^+^**c**, we can then write

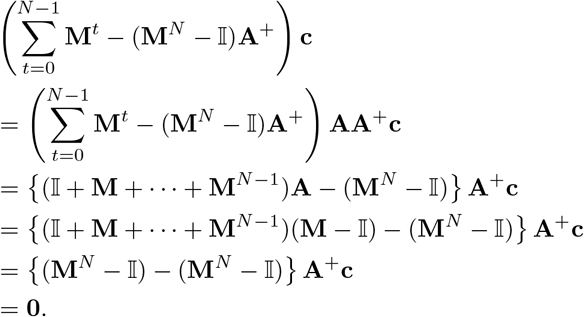

Note that **AA**^+^ is a projection matrix because it is a property of the Moore-Penrose generalised inverse that **AA**^+^**A** = **A**, and therefore that (**AA**^+^)^2^ = **AA**^+^**AA**^+^ = **AA**^+^. Moreover, **AA**^+^ is a projector onto the column space of **A**, because **AA**^+^**A** = **A**. So our desired Equation 30 holds if **c** is in the column space of **A**.

A state in a Markov chain is defined as *recurrent* if the probability of returning to this state at some future iteration is 1; otherwise it is *transient*. The set *S* of states for the Markov chain decomposes into the set of transient states, *T*, and recurrent states, *C*, ie *S* = *T* ∪ *C*. It is known that the set *C* is closed, meaning that once a Markov chain enters *C*, it does not leave it. In health economics, the most common type of recurrent state is an absorbing state (a single state that is never left), often representing ‘dead’. It is natural to assume that costs are zero for recurrent states so that sums of costs for cohorts over time converge; otherwise, one would be assuming some form of cost-accruing immortality. As well as being a natural assumption, we now show that zero costs for recurrent states ensures that **c** is in the column space of **A**, and therefore allows us to use the Moore-Penrose inverse.

First, note that we can reorder states to bring any Markov chain transition matrix into the canonical form (i.e. all recurrent states first, and then afterwards any transient states). We can write this matrix in block form as

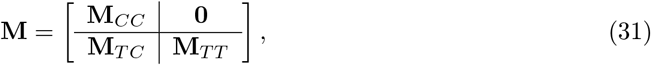

which makes clear how starting points in *T* eventually end up in *C*. No movement from *C* to *T* is possible because the probabilities associated with these transitions (which appear in the upper right block) are all zero.

While **M**_*CC*_ must itself be a stochastic matrix (i.e. the rows sum to 1), **M**_*T T*_ is not; the sums along its rows are *less* than 1, reflecting the ‘leakage’ from *T* to *C*. We can write 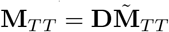 with **D** diagonal and **D**_*ii*_ = Σ_*j*∈*T*_ *p*_*ij*_ *<* 1 the probability of remaining in *T* from state *i* in the next iteration, and the stochastic matrix 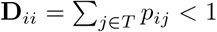 defined by dividing the *i*-th row of **M**_*T T*_ by **D**_*ii*_. Since the maximum absolute eigenvalue of a stochastic matrix is 1 and the eigenvalues of **D** are *<* 1, the matrix **M**_*T T*_ has absolute eigenvalues *<* 1.

Note that **A** has the same block structure as **M** in Equation 31, i.e.

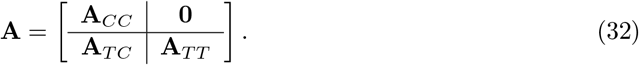

Because **M**_*CC*_ is a stochastic matrix, **A**_*CC*_ = (**M**_*CC*_ − 𝕀) has rows that sum to zero. This means that it must have at least one column that is a linear combination of the remaining columns, and is therefore singular (i.e. not invertible). In contrast, because the eigenvalues of **M**_*T T*_ are *<* 1, **A**_*T T*_ = (**M**_*T T*_ − 𝕀) is non-singular and therefore full rank. This implies that for any **c** that is zero on *C*, i.e.,

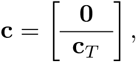

then **c** is in the column space of **A** since **0** is in the column space of all matrices, and **c**_*T*_ is in the column space of **A**_*T T*_. To see why **c**_*T*_ is in the column space of **A**_*T T*_, recall that the column space of an *n × n* non-singular matrix is R*n*, and therefore includes all *n*-dimensional vectors.

Thus, we have shown that if **c** is zero for recurrent states, this is a sufficient condition to allow us to use Equation 30.

## B Appendix: Proof that Simpson’s 1/3 rule and the second-order Gaussian quadrature are equivalent

Our second order approximation for costs is

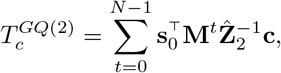

where 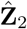 is given by

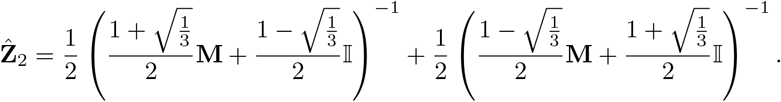

Using standard manipulations, one can show that

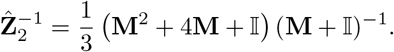

Note we can also write this as

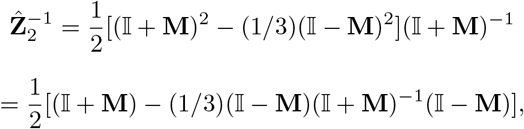

since (𝕀 + **M**)^−1^ commutes with (𝕀 − **M**).

Defining

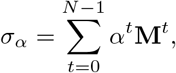

note that

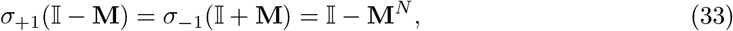

with the last equality holding for even *N*, and also that

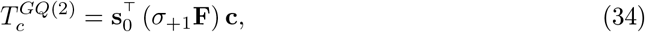

where we can expand the parenthesis in Equation 34 using Equation 33 to find

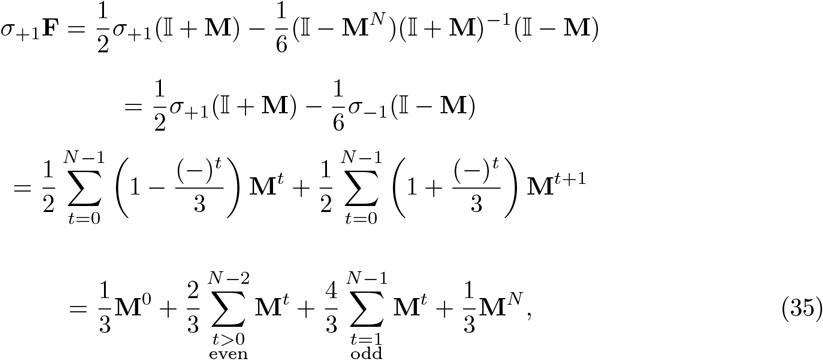

since

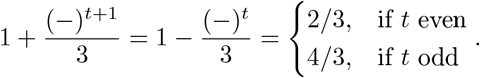

Substituting Equation 35 back for the parenthesis in Equation 34 proves the claimed equivalence of GQ2 and Simpson’s 1/3 rule.

## Supplemental Document

### S1 Impact of time horizon on model error

As a scenario analysis we have varied time horizon of the model and analyse the impact on absolute error between continuous-time and discrete-time model adjusted with different cycle correction methods. Figure S1 presents the analysis result. It can be seen that the higher order GQ method outperformed other existing methods at every instance.

**Figure S1:**
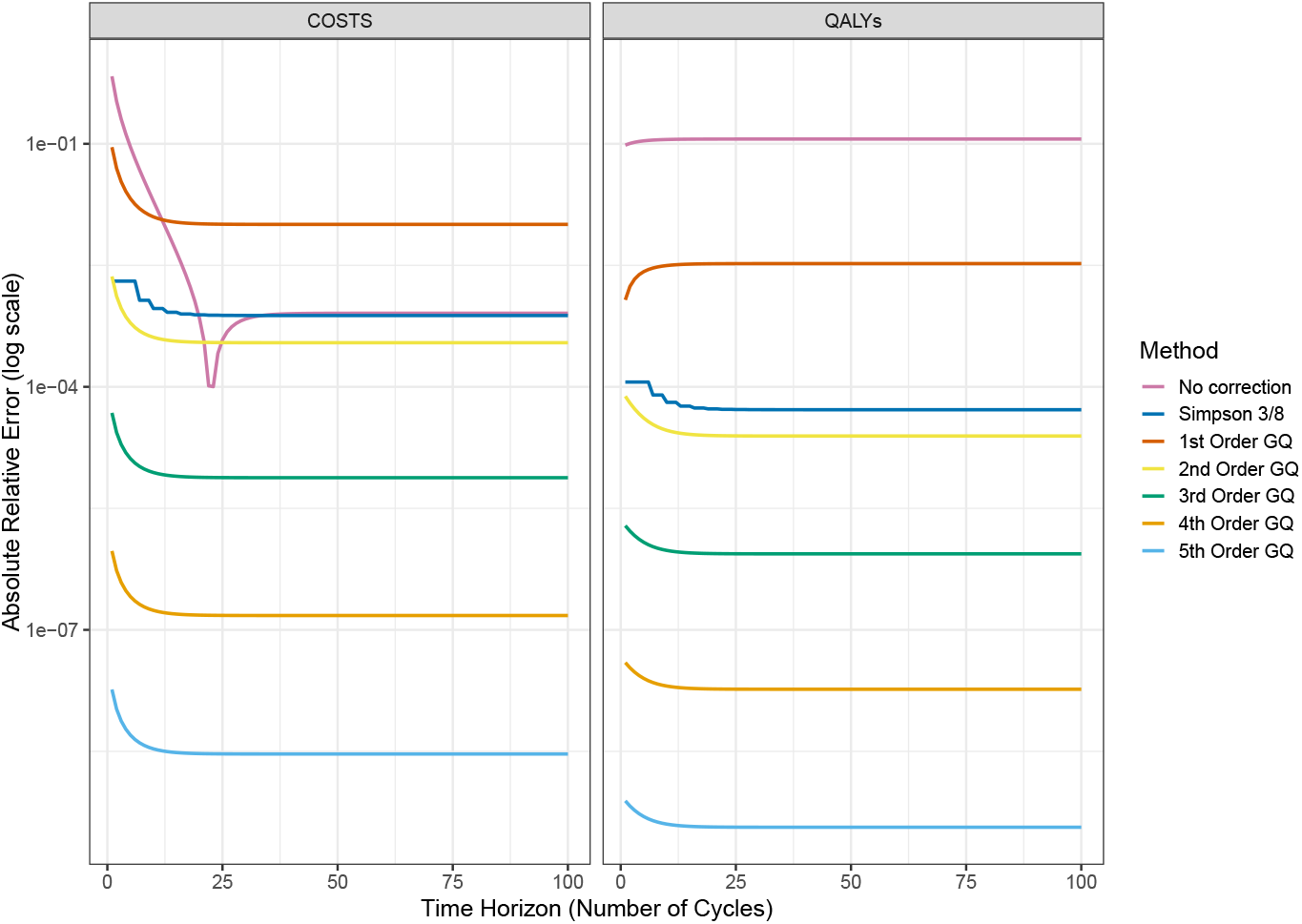
Impact of time horizon on absolute error between continuous- and discrete-time model using different approximation methods. 1st order GQ is same as HCC and trapezoidal and 2nd order GQ is same as Simpson 1/3. Clearly higher order GQ method has lowest error compared to the other existing methods. Simpson 3/8 method was run only for time horizons which were multiple of 3. Abbreviations: GQ: Gaussian Quadrature; HCC: Half-Cycle Correction; QALY: Quality Adjusted Life-Years.

### S2 Discounted results

Table S1 shows the discounted results. 3.5 % annual discount rate was considered for both costs and QALYs.

**Table S1:** Comparative analysis discounted result per person based on different methods. Absolute error is presented in increasing order of error. Discounting was performed before cycle correction. 5th order GQ method has the absolute minimum error for both costs and QALYs and closest to continuous model. 1st order GQ is same as HCC and trapezoidal and 2nd order GQ is same as Simpson 1/3. NMB is with respect to continuous time model with willingness-to-pay of £30,000. Abbreviations: GQ: Gaussian Quadrature; HCC: Half-Cycle Correction; NMB: Net Monetary Benefit; QALY: Quality Adjusted Life-Years

| Correction Method | Costs | Absolute Error | QALYs | Absolute Error | NMB |
| --- | --- | --- | --- | --- | --- |
| Continuous time model | £190.5293 | - | 3.73378 | - | - |
| 5th order GQ method | £190.5293 | 8.48e-09 | 3.73378 | 1.26e-07 | -£0.1341 |
| 4th order GQ method | £190.5293 | 2.74e-07 | 3.73378 | 1.57e-07 | -£0.1375 |
| 3rd order GQ method | £190.5270 | 1.20e-05 | 3.73377 | 1.39e-06 | -£0.2739 |
| 2nd order GQ method | £190.4363 | 4.88e-04 | 3.73368 | 2.62e-05 | -£2.9653 |
| Simpson's 3/8 method | £190.3289 | 1.05e-03 | 3.73357 | 5.40e-05 | -£5.9629 |
| 1st order GQ method | £188.2323 | 1.21e-02 | 3.75069 | 4.53e-03 | £509.5421 |
| No correction performed | £190.7323 | 1.07e-03 | 4.22569 | 1.32e-01 | £14,757.0421 |

### S3 Where is 5th order GQ less likely to be the most accurate approximation?

Figure S2 shows which parameter values are less likely to result in 5th order GQ being the best approximation method. Figure S3 explores interactions effects, ie. whether certain pairwise combinations of parameters are less likely to result in 5th order GQ being the best approximation method.

**Figure S2:**
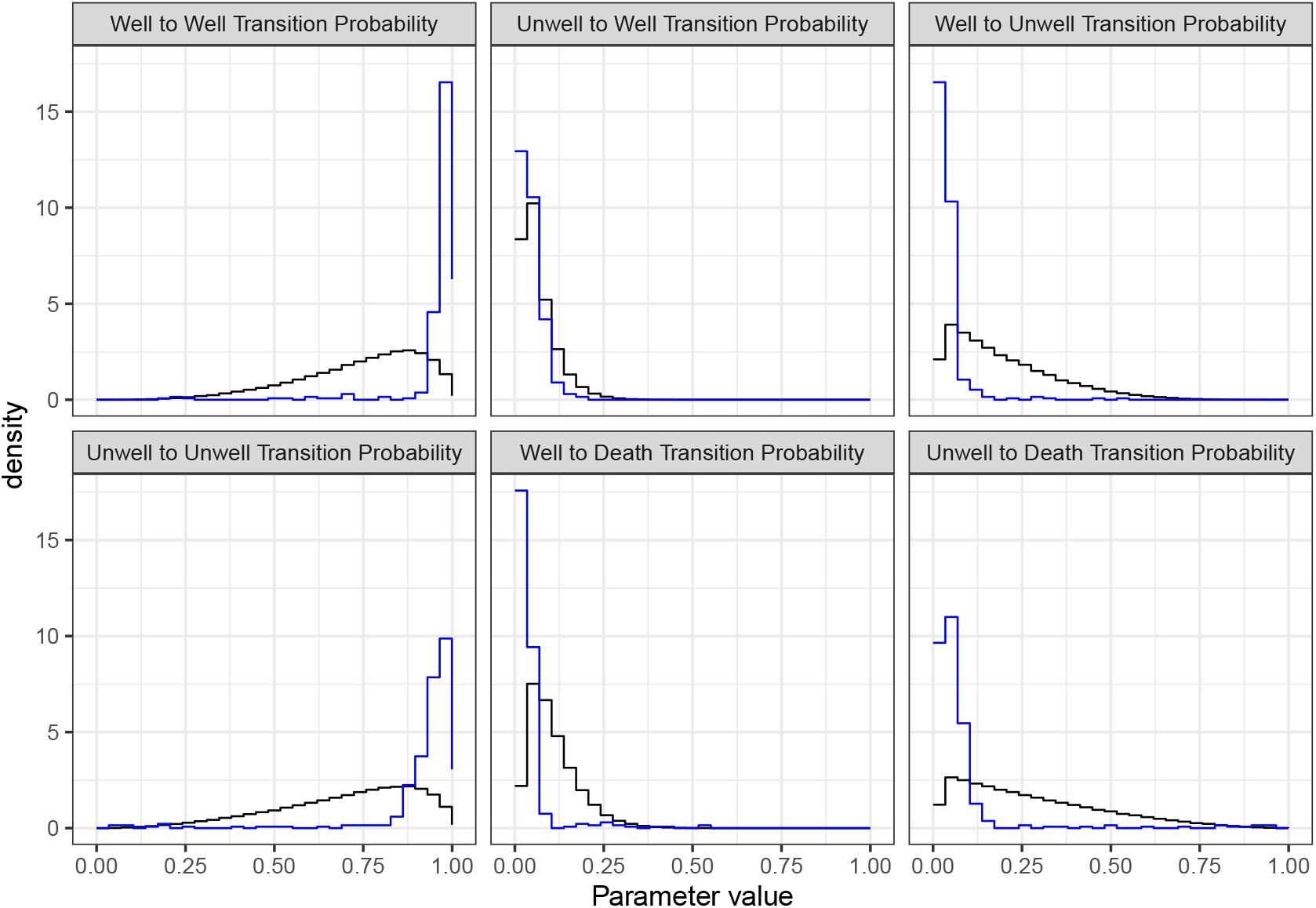
Parameter histograms when 5th order GQ is not best in 100,000 simulations (blue), compared to sampled parameters (black).

**Figure S3:**
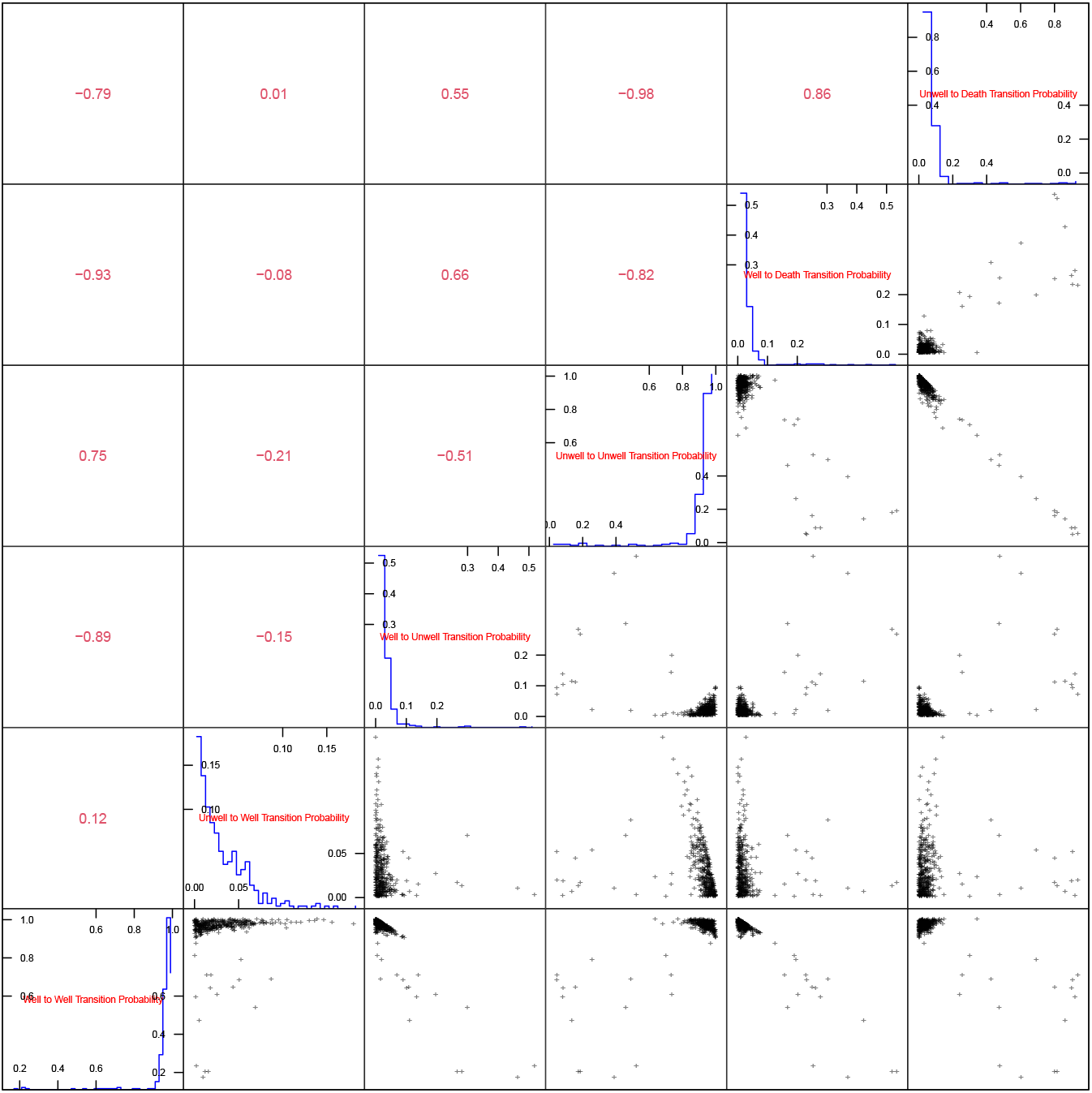
Pairs plot of parameters for which 5th order GQ is not best in 100,000 simulations.

### S4 Expressions for 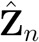

Given an unadjusted discrete-time Markov model

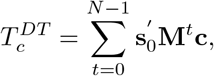

the *n*-th order Gaussian quadrature approximation to the continuous-time model output is given by

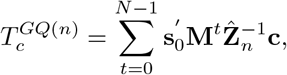

where 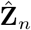 is defined as

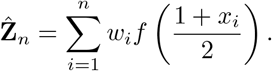

The function *f*(·) is defined as

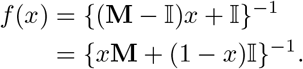

Locations *{x*_*i*_*}* and weights *{w*_*i*_*}* for the first 5 orders of Gaussian-Legendre quadrature are

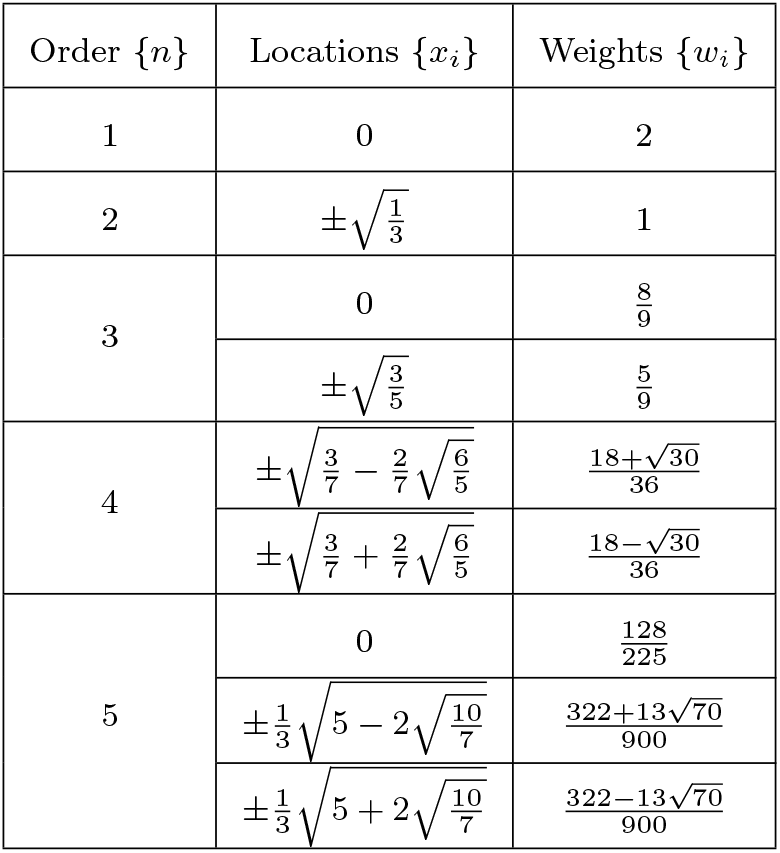

This leads the following expressions for 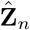 for *n* = 1, …, 5:

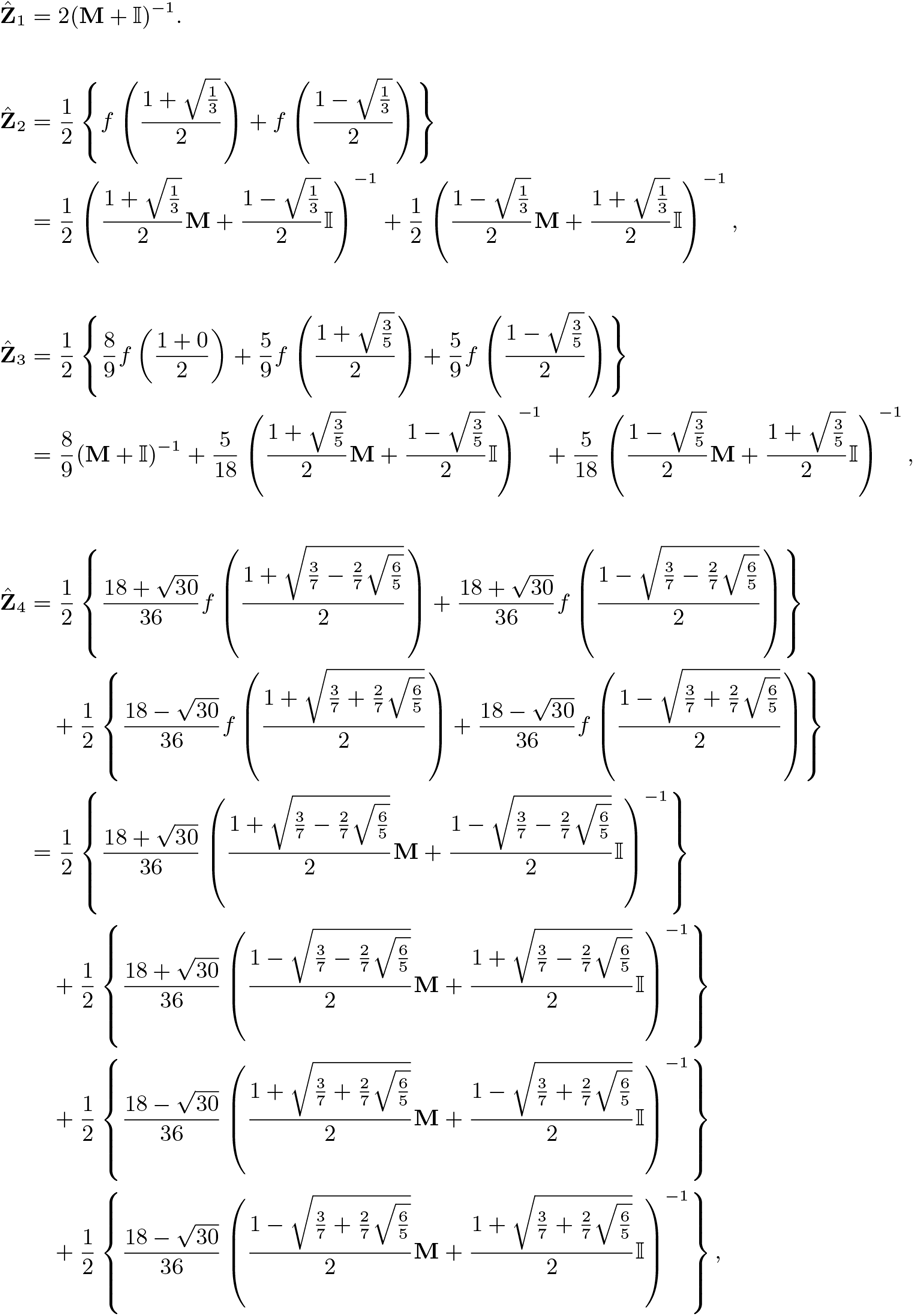

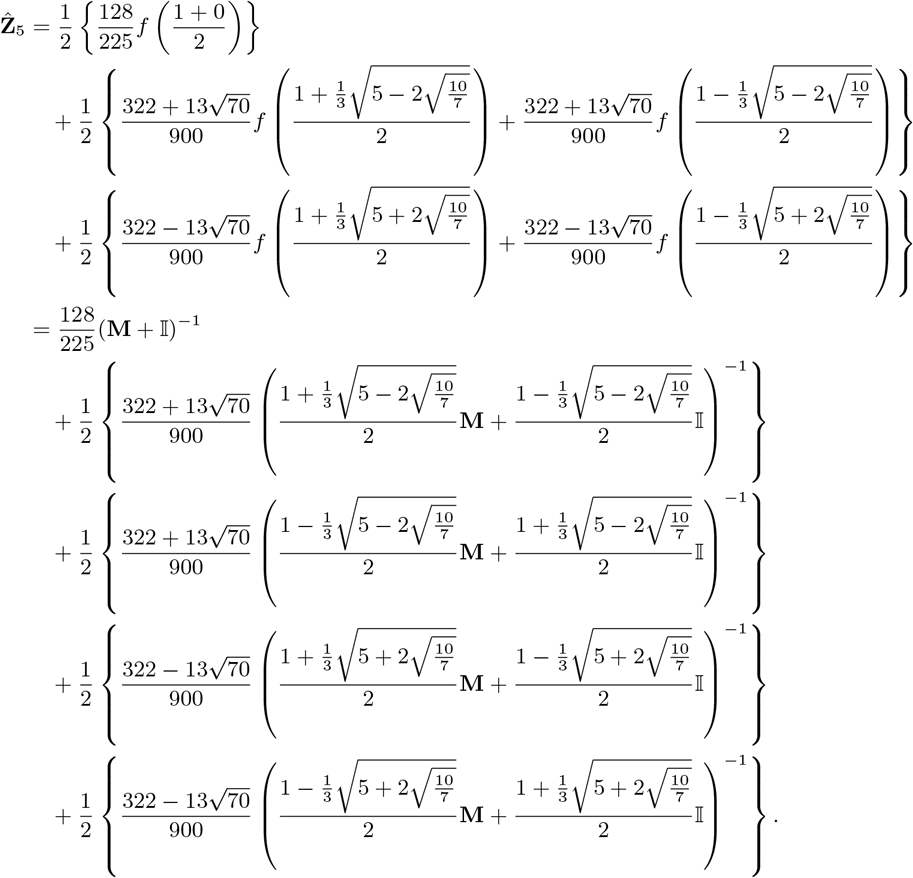

### S5 Distribution of input parameters

Figure S4 presents distribution of input parameters of the model considered for probabilistic sensitivity analysis.

**Figure S4:**
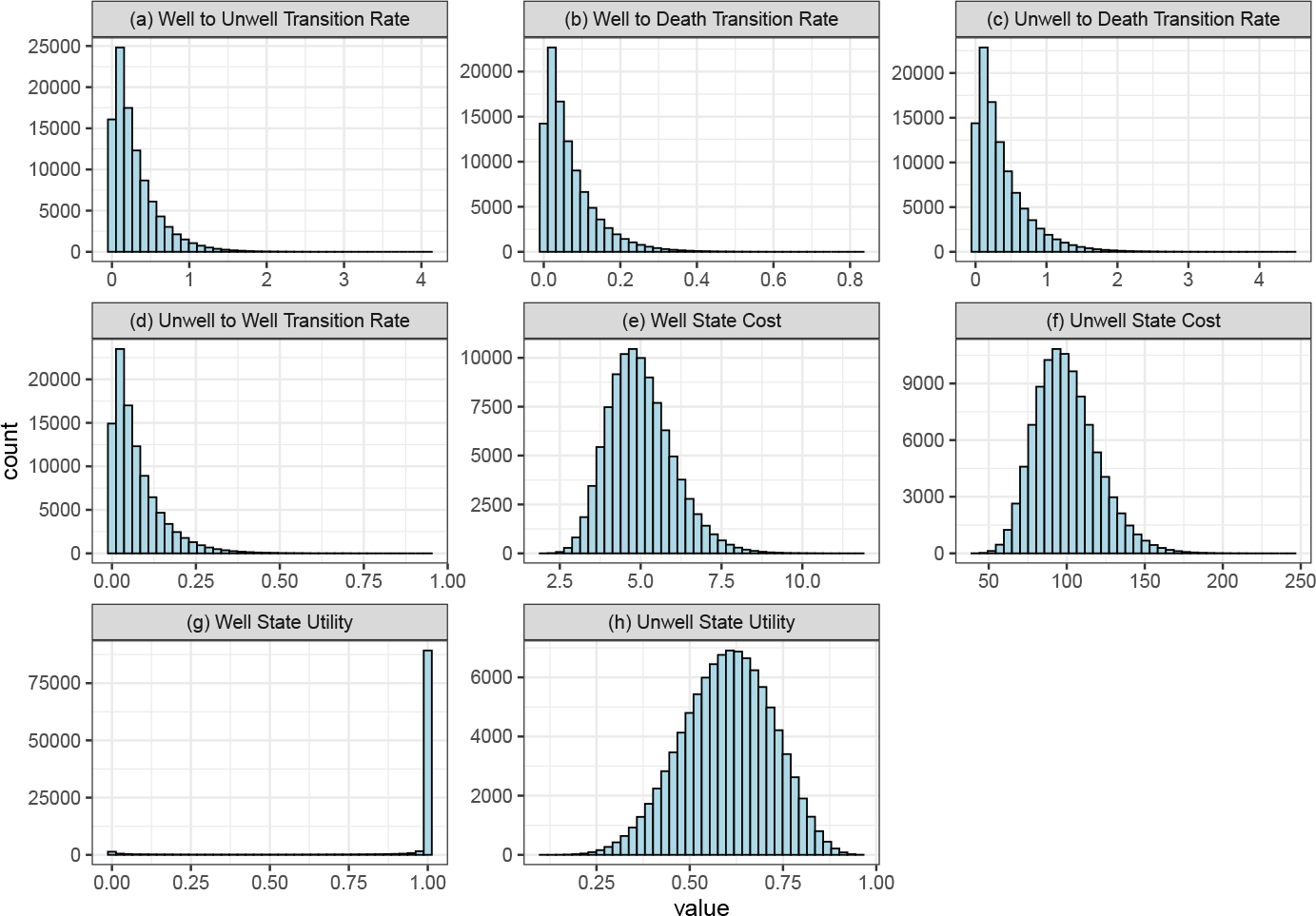
Distribution used in PSA for each model input parameters (100,000 samples were used).Abbreviation: PSA: Probabilistic Sensitivity Analysis

